# Clinical effectiveness of SARS-CoV-2 booster vaccine against Omicron infection in residents and staff of Long-Term Care Facilities: a prospective cohort study (VIVALDI)

**DOI:** 10.1101/2022.08.08.22278532

**Authors:** Oliver Stirrup, Madhumita Shrotri, Natalie L. Adams, Maria Krutikov, Hadjer Nacer-Laidi, Borscha Azmi, Tom Palmer, Christopher Fuller, Aidan Irwin-Singer, Verity Baynton, Gokhan Tut, Paul Moss, Andrew Hayward, Andrew Copas, Laura Shallcross

## Abstract

**Background:** Successive SARS-CoV-2 variants have caused severe disease in long-term care facility (LTCF) residents. Primary vaccination provides strong short-term protection, but data are limited on duration of protection following booster vaccines, particularly against the Omicron variant. We investigated effectiveness of booster vaccination against infections, hospitalisations and deaths among LTCF residents and staff in England.

**Methods:** We included residents and staff of LTCFs within the VIVALDI study (ISRCTN 14447421) who underwent routine, asymptomatic testing (December 12 2021-March 31 2022). Cox regression was used to estimate relative hazards of SARS-CoV-2 infection, and associated hospitalisation and death at 0-13, 14-48, 49-83 and 84 days after dose 3 of SARS-CoV-2 vaccination compared to 2 doses (after 84+ days), stratified by previous SARS-CoV-2 infection and adjusting for age, sex, LTCF capacity and local SARS-CoV-2 incidence.

**Results:** 14175 residents and 19973 staff were included. In residents without prior SARS-CoV-2 infection, infection risk was reduced 0-83 days after first booster, but no protection was apparent after 84 days. Additional protection following booster vaccination waned, but was still present at 84+ days for COVID-associated hospitalisation (aHR: 0.47, 0.24-0.89) and death (aHR: 0.37, 0.21-0.62). Most residents (64.4%) had received primary course of AstraZeneca, but this did not impact on pre- or post-booster risks. Staff showed a similar pattern of waning booster effectiveness against infection, with few hospitalisations and no deaths.

**Conclusions:** Booster vaccination provides sustained protection against severe outcomes following infection with the Omicron variant, but no protection against infection from 3 months onwards. Ongoing surveillance for SARS-CoV-2 in LTCFs is crucial.

**Summary:** The COVID-19 pandemic has severely impacted residents in long-term care facilities (LTCFs). Booster vaccination provides sustained moderate protection against severe outcomes, but no protection against infection was apparent from around 3 months onwards. Ongoing surveillance in LTCFs is crucial.

## Introduction

The disproportionate impact of COVID-19 on long-term care facilities (LTCFs) has been extensively documented, both in terms of direct effects of morbidity and mortality, and indirect consequences of reduced access to healthcare, services and social interactions[1]. Likely reasons for this include the closed-setting environment of LTCFs, impaired immune responses due to aging and high levels of comorbidity[2]. As such, LTCF residents and staff were prioritised for vaccination against SARS-CoV-2.

Vaccination programmes in UK LTCFs commenced on 8 December 2020[3, 4], with primary series delivery of homologous prime-boost with either BNT162b2 (Pfizer) or ChAdOx1 (AstraZeneca) using an 8-12 week dose interval. LTCF residents and staff were prioritised for additional booster vaccination (third dose) from 14 September 2021 with a fourth dose for adults aged 75 years and over and older residents in LTCFs from Spring 2022[5] based on evidence that protection against severe disease waned from six months following primary vaccination in older adults[6], and concerns about the impact of new variants.

We previously reported high level of short-term protection against infection and severe clinical outcomes following primary course vaccination in LTCF residents and staff, whilst Alpha and Delta variants were dominant[7]. However, we observed substantial waning of protection against infection in staff and against all outcomes (infection, hospital admission, death) in residents from 12 weeks following second dose, which was restored following a booster dose[7]. Substantially increased short term protection against symptomatic disease, hospitalisation and death was also seen in adults >50 years following a third dose booster vaccination during high prevalence of Delta variant, with limited waning of protection after 10 weeks [8]. Data on booster vaccination effectiveness against Omicron in LTCFs are limited, particularly following a primary course of AstraZeneca. However, three vaccine doses have been reported to offer high levels of protection against hospitalisation with the Omicron variant in community-dwelling adults aged 75 years and over, with minimal waning 2-3 months post-vaccination[9].

The aim of this study was to evaluate the effectiveness of third and fourth dose booster vaccination against infection, hospitalisations and death amongst staff and residents of LTCFs in England, from when the Omicron variant became dominant until the end of the asymptomatic testing programme in LTCF residents (December 12 2021 to March 31 2022).

## Methods

### Study design and setting

VIVALDI is a prospective cohort study investigating SARS-CoV-2 in residents and staff in LTCFs in England and is described in detail elsewhere[10]. In the analysis period, following national guidelines, residents were undergoing monthly routine polymerase chain reaction (PCR) testing while staff were undergoing weekly testing using a combination of PCR and lateral flow devices (LFD). We did not require PCR confirmation of positive LFD results in our analyses. Individuals with a positive PCR test were not routinely re-tested for 90 days unless they developed new COVID-19 symptoms[11].

The analysis period is defined from December 12, 2021, when the S-gene target failure (SGTF) marker for Omicron (BA.1) was first detected in the dataset[12], to 31 March 2022, when asymptomatic testing in residents ended[13]. Individuals were eligible for inclusion if they had at least one PCR or LFD result available within the analysis period. We excluded individuals who had not received at least two vaccine doses at the start of the analysis period.

Ethical approval was obtained from South Central-Hampshire B Research Ethics Committee (20/SC/0238). The legal basis to access data from staff and residents without informed consent was provided by Regulation 3(4) of the Health Service (Control of Patient Information) Regulations 2002 (COPI)[14].

### Data extraction and linkage

We retrieved all PCR results and available LFD results from routine symptomatic and asymptomatic testing in LTCFs, and positive PCR results from staff and residents who underwent clinical testing in hospitals through the COVID-19 Datastore[15]. Test results, vaccination, hospitalisation and deaths data were linked to study participants using pseudo-identifiers based on individuals’ unique National Health Service (NHS) numbers[7]. COVID-19 hospitalisation was defined as an admission within 14 days of a positive PCR or LFD test for SARS-CoV-2, or admission with positive test on the same or subsequent day. COVID-19 death was defined as within 28 days of positive PCR or LFD test.

As previously[7], we linked SARS-CoV2 serological test results for IgG antibodies to the nucleocapsid protein (Abbott ARCHITECT system (Abbott, Maidenhead, UK)) in a subset of participants. We combined positive PCR and LFD results, COVID-19-related hospital admission records, and positive nucleocapsid antibody results before the analysis period into a binary variable indicating evidence of prior SARS-CoV2 exposure.

Care Quality Commission unique location identification (CQC-ID) were used to link residents to LTCFs. Data on bed capacity were retrieved from Capacity Tracker[16], and requested directly from LTCF managers if unavailable. Seven-day rolling rates of SARS-CoV2 incidence at local authority level[17] were used to represent local infection pressure for each LTCF. A data privacy impact assessment was completed for the VIVALDI study and a privacy notice published[18].

### Statistical analysis

We examined individual-level vaccine effectiveness against infection, hospitalisation within 14 days and death within 28 days of positive test, separately for residents aged 65 years and older, and for staff between 18 and 75 years. Individuals were eligible for inclusion if they had complete data on sex and age, had received at least 2 vaccine-doses 12 or more weeks before analysis start date, had at least one PCR or LFD test result recorded within the analysis period and were linked to a LTCF with data on total number of staff and residents. Individuals with third vaccine dose recorded prior to start of official rollout on September 14 2021 were excluded.

We used Cox regression models to derive adjusted hazard ratios (HRs) for the risk of each outcome of interest. Vaccination status was included as a time-varying covariable. The reference category was 2 vaccine doses, with 12 or more weeks (84 days) elapsed from Dose 2. The exposure categories were 0–13, 14–48, 49–83 and 84 or more days following Dose 3, and any time following Dose 4. Individuals could start in the 2-dose vaccinated state and sequentially transition through 3- and 4-dose vaccinated exposure states. Individuals entered the risk period on 12 December 2021, or date of their first recorded PCR/LFD result within VIVALDI if later.

Individuals with positive PCR/LFD result within 30 days prior to 12 December 2021 entered the risk period from the 31st day post-positive test. Individuals exited the risk period at the earliest of: outcome of interest or end of analysis period. For hospitalisation, individuals were additionally censored at 15 days post-positive SARS-CoV-2 test if there was no hospital admission by then, and similarly for mortality after 29 days. Baseline hazard was defined over calendar time. 95% CIs were calculated using robust SEs accounting for dependence of infection events within LTCFs.

The primary analysis was stratified by evidence of SARS-CoV2 infection prior to the risk period. We adjusted for sex (binary variable), age (5-knot restricted cubic spline term), LTCF size expressed as total number of beds (linear term), local SARS-CoV2 incidence expressed as 7-day rolling rate per 100 population. Local incidence was included as a 3-knot restricted cubic spline term with separate coefficients for each calendar month. We also evaluated effect of AstraZeneca vs Pfizer primary vaccination course (as recorded for Dose 2) on each outcome. This effect was estimated separately before and after initial booster dose (Dose 3) and tested jointly using a multivariate Wald test.

We also conducted descriptive analysis of the incidence of new and repeat SARS-CoV-2 infections in LTCF staff and residents from October 2020 onwards (when regular testing had been implemented) to evaluate whether the Omicron variant was associated with a rise in reinfections. Repeat infections were considered to be any positive PCR/LFD test recorded over 30 days after previous positive test. Participants were considered to be under follow-up from first recorded PCR/LFD test until 90 days following their last recorded test.

All statistical analyses were conducted using STATA 17.0.

## Results

14175 residents and 19793 staff from 328 LTCFs (Table S1) were included in the analysis (Figure 1). 2680 residents (18.9%) and 2488 staff (12.6%) had recorded evidence of SARS-CoV-2 infection prior to the analysis period (Table 1). 60955 PCR (mean±SD per month, 1.92±2.34) and 27539 LFD (0.76±2.36 per month) tests for residents, and 80502 PCR (1.62±2.02 per month) and 281983 LFD (5.16±6.37 per month) tests for staff were included.

**Figure 1.**
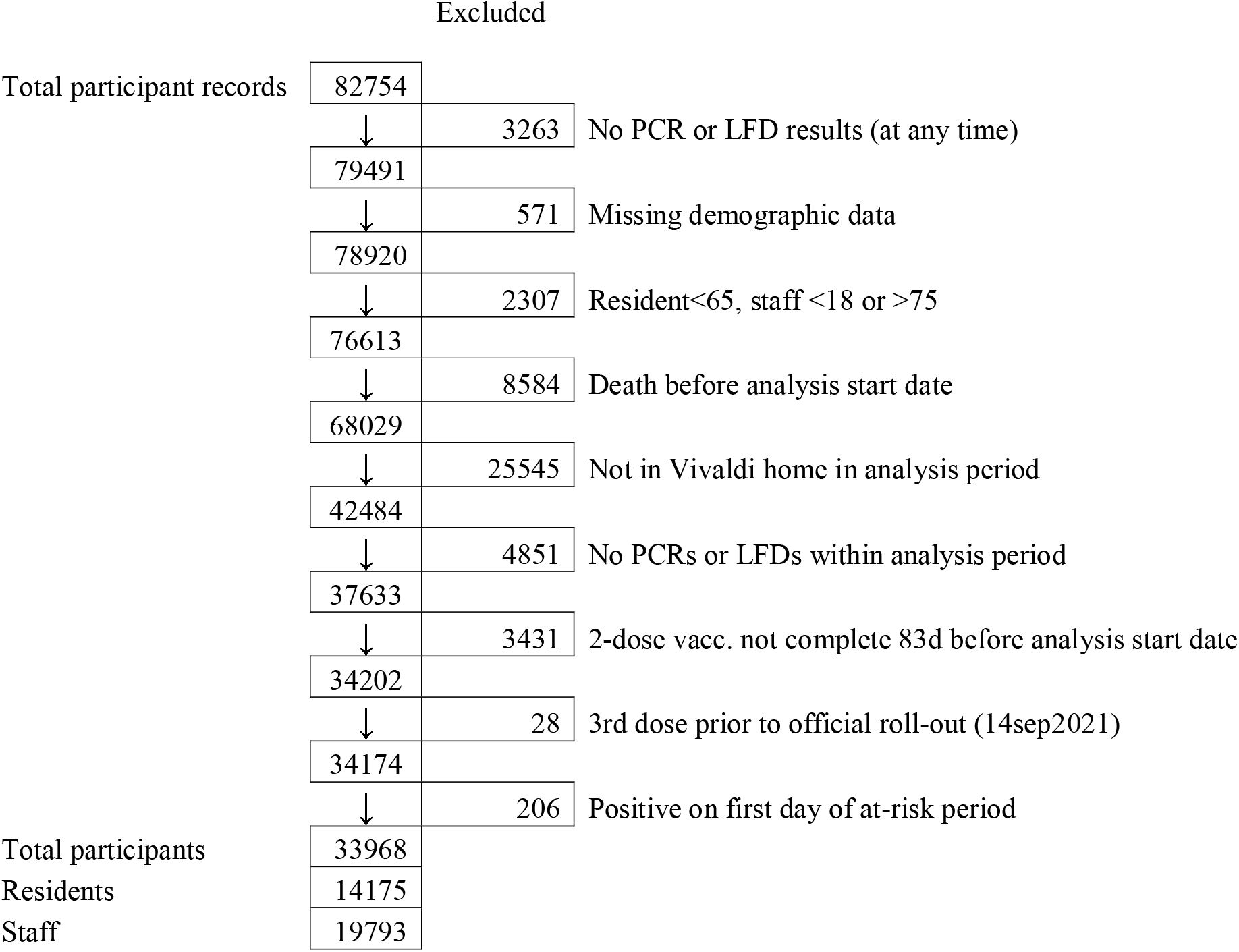
Flow chart for inclusion in the analysis of booster vaccine effectiveness.

**Table 1.**
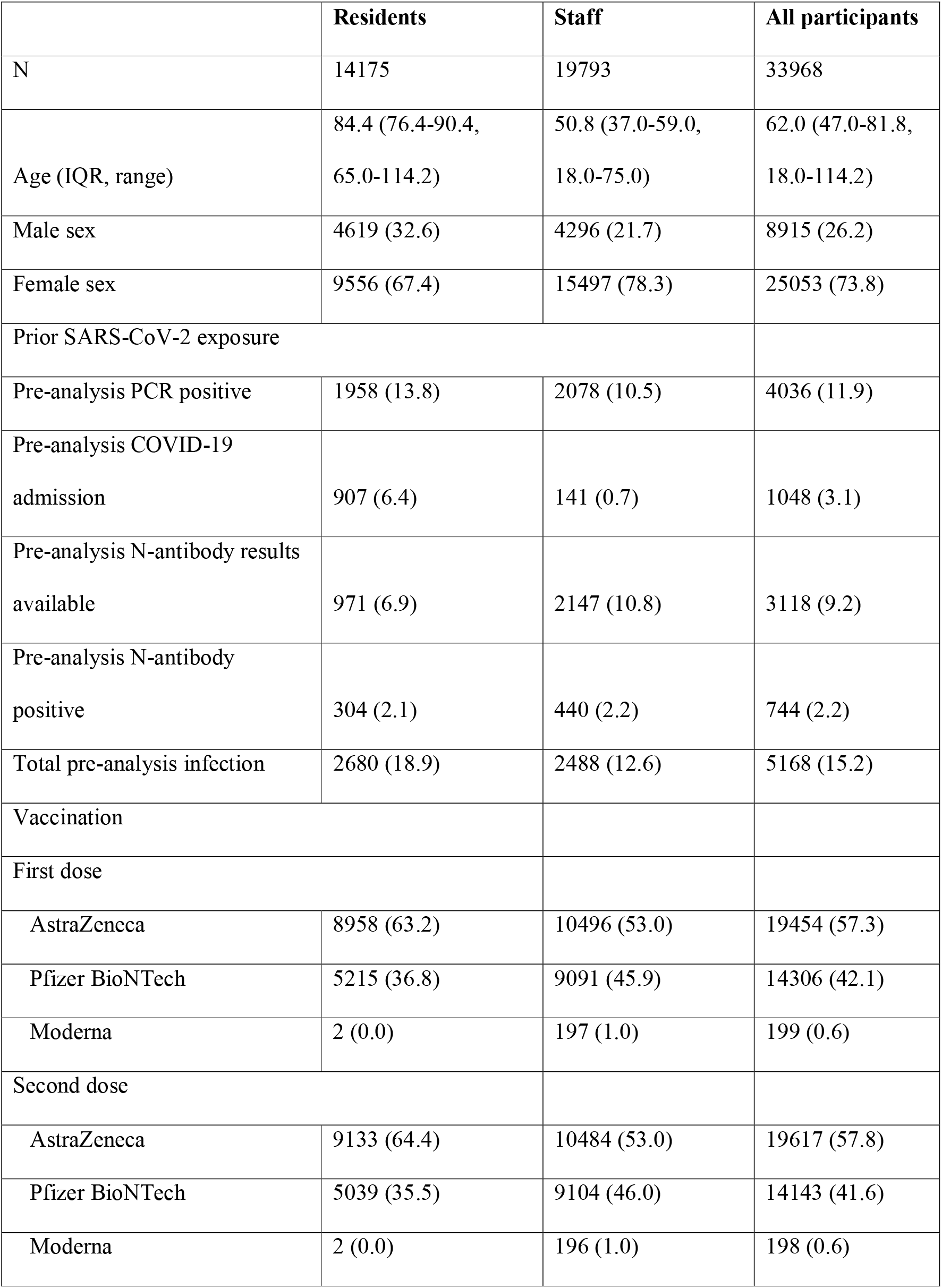
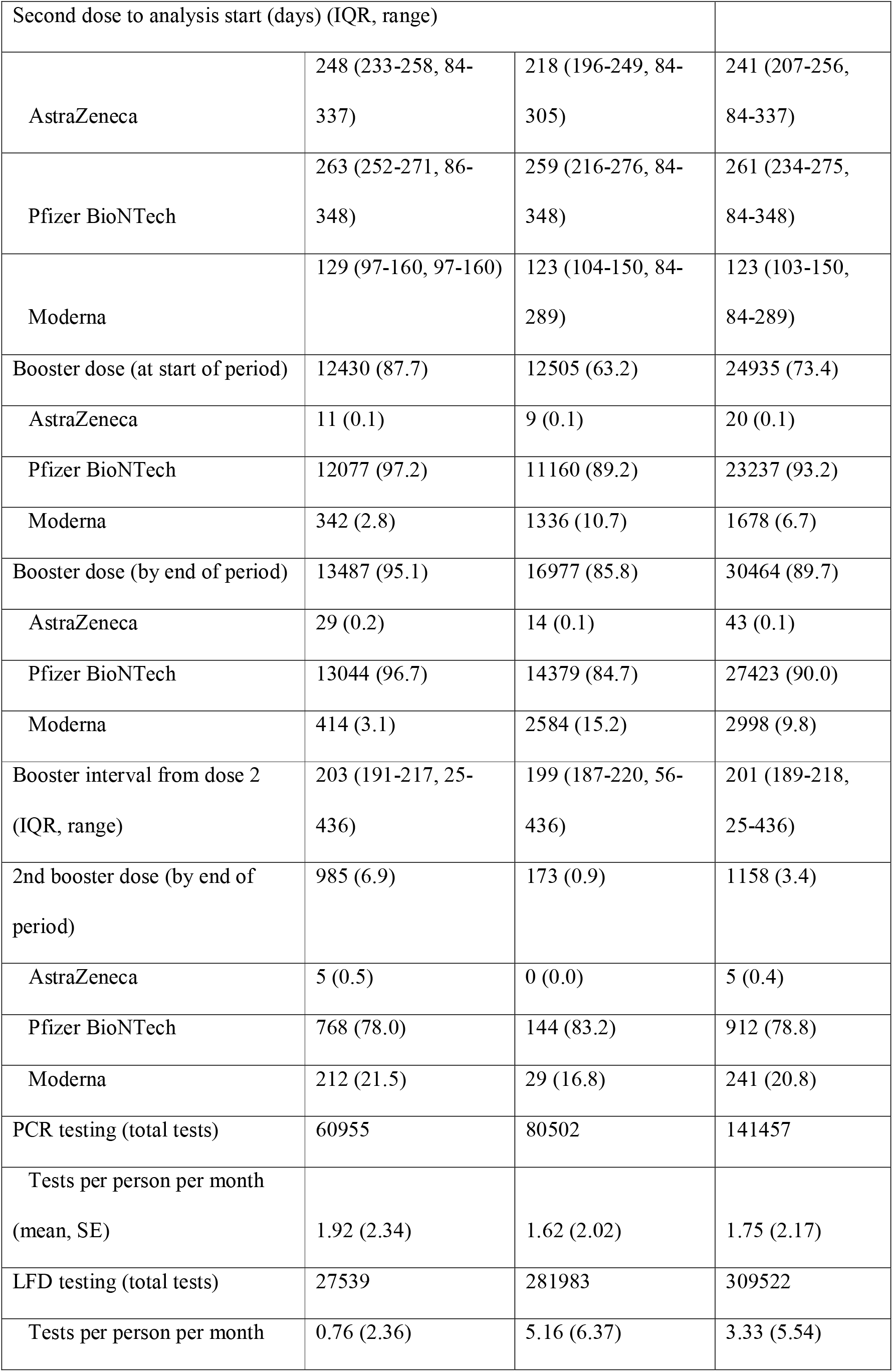
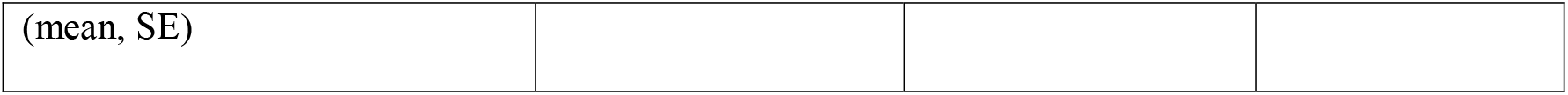
Characteristics of residents and staff included in the analysis of booster vaccine effectiveness.

12430 (87.7%) residents and 12505 (63.2%) staff had received a booster vaccination dose prior to the analysis period, and 13487 (95.1%) residents and 16977 (85.8%) staff by the end. First booster doses were Pfizer in the majority (residents n=13044, 96.7%; staff n=14379, 84.7%), with Moderna used in a small number (residents n=414, 3.1%; staff n=2584, 15.2%). Second boosters (fourth vaccine) had been received by 6.9% of residents and 0.9% of staff by the end of the analysis period.

### Infection

In residents without known prior SARS-CoV-2 infection, there was reduced risk of SARS-CoV-2 infection in the periods 0-13 days (HR 0.48, 95% CI 0.31-0.73), 14-48 days (HR 0.28, 0.20-0.38) and 49-83 days (0.31, 0.24-0.40) after first booster vaccine dose, relative to 2-dose vaccination (Table 2). However, no protection was apparent at 84+ days following booster vaccination (1.13, 0.93-1.39). Residents with known infection prior to the analysis period were at reduced risk of new infection relative to those without prior infection (HR 0.53, 0.37-0.77), and within this group further protection following booster vaccination followed a similar pattern to that observed in infection-naïve residents. Infection rates were lower after second booster doses, but HR estimates were wide due to limited follow-up time. Additional adjustment for type of primary course before and after booster dose did not improve model fit (P=0.46).

**Table 2.** Crude event rates and adjusted hazard ratios against PCR or LFD-positive SARS-CoV2 infections, hospitalisation within 14 days and deaths within 28 days of a positive PCR or LFD test for LTCF residents, by prior SARS-CoV2 exposure, and vaccination status

| **SARS-CoV-2 infections** |  |  |  |  |  |  |
| --- | --- | --- | --- | --- | --- | --- |
| **Prior SARS-CoV-2 exposure** | **Vaccination status** | **Person-days** | **Infections** | **IR /1000pd** | **HR (95% CI)\*** | **HR (95% CI)\*** |
| Unexposed | D2 84+d | 50497 | 215 | 4.26 | Ref. |  |
|  | D3 0-13d | 10416 | 41 | 3.94 | 0.48 (0.31-0.73) |  |
|  | D3 14-48d | 75643 | 142 | 1.88 | 0.28 (0.20-0.38) |  |
|  | D3 49-83d | 245757 | 538 | 2.19 | 0.31 (0.24-0.40) |  |
|  | D3 84+d | 571574 | 1636 | 2.86 | 1.13 (0.93-1.39) |  |
|  | D4 0+d | 6119 | 14 | 2.29 | 0.93 (0.40-2.13) |  |
| Exposed | D2 84+d | 16860 | 31 | 1.84 | 0.53 (0.37-0.77) | Ref. |
|  | D3 0-13d | 3138 | 5 | 1.59 |  | 0.33 (0.12-0.87) |
|  | D3 14-48d | 18935 | 12 | 0.63 |  | 0.17 (0.09-0.33) |
|  | D3 49-83d | 63290 | 88 | 1.39 |  | 0.35 (0.22-0.57) |
|  | D3 84+d | 149787 | 252 | 1.68 |  | 1.41 (0.93-2.13) |
|  | D4 0+d | 1477 | 1 | 0.68 |  | 0.46 (0.06-3.37) |
| **SARS-CoV-2 hospitalisations** |  |  |  |  |  |  |
| **Prior SARS-CoV-2 exposure** | **Vaccination status** | **Person-days** | **Hosp.** | **IR /1000pd** | **HR (95% CI)\*** | **HR (95% CI) \*** |
| Unexposed | D2 84+d | 53381 | 15 | 0.28 | Ref. |  |
|  | D3 0-13d | 10687 | 2 | 0.19 | 0.27 (0.05-1.34) |  |
|  | D3 14-48d | 77372 | 7 | 0.09 | 0.19 (0.07-0.52) |  |
|  | D3 49-83d | 251742 | 21 | 0.08 | 0.15 (0.07-0.32) |  |
|  | D3 84+d | 594878 | 42 | 0.07 | 0.47 (0.24-0.89) |  |
|  | D4 0+d | 6193 | 0 | 0 | — |  |
| Exposed | D2 84+d | 17301 | 0 | 0 | — | Ref. |
|  | D3 0-13d | 3201 | 0 | 0 |  | — |
|  | D3 14-48d | 19086 | 0 | 0 |  | — |
|  | D3 49-83d | 64263 | 1 | 0.02 |  | — |
|  | D3 84+d | 153592 | 2 | 0.01 |  | — |
|  | D4 0+d | 1479 | 0 | 0 |  | — |
| **SARS-CoV-2 mortality** |  |  |  |  |  |  |
| **Prior SARS-CoV-2 exposure** | **Vaccination status** | **Person-days** | **Deaths (w. 28d)** | **IR /1000pd** | **HR (95% CI)\*** | **HR (95% CI) \*** |
| Unexposed | D2 84+d | 55752 | 23 | 0.41 | Ref. |  |
|  | D3 0-13d | 10698 | 1 | 0.09 | 0.14 (0.02-1.04) |  |
|  | D3 14-48d | 78442 | 6 | 0.08 | 0.12 (0.04-0.34) |  |
|  | D3 49-83d | 254513 | 18 | 0.07 | 0.11 (0.05-0.23) |  |
|  | D3 84+d | 615354 | 83 | 0.13 | 0.37 (0.21-0.62) |  |
|  | D4 0+d | 6282 | 0 | 0 | — |  |
| Exposed | D2 84+d | 17637 | 0 | 0 | — | Ref. |
|  | D3 0-13d | 3210 | 0 | 0 |  | — |
|  | D3 14-48d | 19191 | 1 | 0.05 |  | — |
|  | D3 49-83d | 64645 | 0 | 0 |  | — |
|  | D3 84+d | 156882 | 13 | 0.08 |  | — |
|  | D4 0+d | 1479 | 0 | 0 |  | — |

\*HR values in the two columns are from mathematically identical statistical models, but HRs in right-hand column are expressed relative to 'D2 84+d' vaccine status in individuals with prior SARS-CoV-2 infection.

A protective effect of first booster vaccine dose was also seen in staff, although no protection was apparent by 84+ days (1.20, 1.09-1.34) (Table 3). However, prior infection was not associated with any reduction in risk of new SARS-CoV-2 infection in this group (HR 1.04, 0.89-1.22) and a similar pattern of protection from infection following first booster dose was observed in this group.

**Table 3.** Crude event rates and adjusted hazard ratios against PCR or LFD-positive SARS-CoV2 infections, hospitalisation within 14 days and deaths within 28 days of a positive PCR or LFD test for LTCF staff, by prior SARS-CoV2 exposure, and vaccination status

| **SARS-CoV-2 infections** |  |  |  |  |  |  |
| --- | --- | --- | --- | --- | --- | --- |
| **Prior SARS-CoV-2 exposure** | **Vaccination status** | **Person-days** | **Infections** | **IR /1000pd** | **HR (95% CI)\*** | **HR (95% CI) \*** |
| Unexposed | D2 84+d | 247325 | 943 | 3.81 | Ref. |  |
|  | D3 0-13d | 51795 | 147 | 2.84 | 0.28 (0.23-0.35) |  |
|  | D3 14-48d | 226481 | 445 | 1.96 | 0.31 (0.27-0.36) |  |
|  | D3 49-83d | 363113 | 753 | 2.07 | 0.45 (0.40-0.50) |  |
|  | D3 84+d | 646022 | 1750 | 2.71 | 1.20 (1.08-1.34) |  |
|  | D4 0+d | 4676 | 6 | 1.28 | 0.77 (0.33-1.75) |  |
| Exposed | D2 84+d | 43850 | 165 | 3.76 | 1.04 (0.89-1.22) | Ref. |
|  | D3 0-13d | 6451 | 10 | 1.55 |  | 0.12 (0.06-0.24) |
|  | D3 14-48d | 25190 | 36 | 1.43 |  | 0.22 (0.15-0.32) |
|  | D3 49-83d | 49207 | 119 | 2.42 |  | 0.42 (0.33-0.53) |
|  | D3 84+d | 108378 | 307 | 2.83 |  | 1.20 (0.99-1.45) |
|  | D4 0+d | 418 | 0 | 0 |  | — |
| **SARS-CoV-2 hospitalisations** |  |  |  |  |  |  |
| **Prior SARS-CoV-2 exposure** | **Vaccination status** | **Person-days** | **Hosp.** | **IR /1000pd** | **HR (95% CI)\*** | **HR (95% CI) \*** |
| Unexposed | D2 84+d | 260532 | 2 | 0.01 | — |  |
|  | D3 0-13d | 52898 | 1 | 0.02 | — |  |
|  | D3 14-48d | 232608 | 3 | 0.01 | — |  |
|  | D3 49-83d | 372655 | 7 | 0.02 | — |  |
|  | D3 84+d | 671523 | 6 | 0.01 | — |  |
|  | D4 0+d | 4731 | 0 | 0 | — |  |
| Exposed | D2 84+d | 46349 | 2 | 0.04 | — | — |
|  | D3 0-13d | 6499 | 0 | 0 |  | — |
|  | D3 14-48d | 25703 | 1 | 0.04 |  | — |
|  | D3 49-83d | 50637 | 1 | 0.02 |  | — |
|  | D3 84+d | 112970 | 2 | 0.02 |  | — |
|  | D4 0+d | 418 | 0 | 0 |  | — |

\*HR values in the two columns are from mathematically identical statistical models, but HRs in right-hand column are expressed relative to ‘D2 84+d’ vaccine status in individuals with prior SARS-CoV-2 infection

Additional adjustment for type of primary course before and after booster dose improved model fit (P=0.03). A primary course of AstraZeneca rather than mRNA-based vaccine (Pfizer or Moderna) was associated with slightly higher risk of infection among staff prior to booster dose (HR 1.12, 0.97-1.28), but not following first booster dose (0.91, 0.83-1.01). Subsequently fitting the Cox model separately by primary series vaccine type suggested marginally stronger additional protection from booster vaccination following a course of AstraZeneca compared to mRNA (Table S2).

### Hospitalisation

In residents without known prior SARS-CoV-2 infection, the first booster dose reduced risk of hospitalisation within 0-13 days of SARS-CoV-2 infection (HR 0.27, 95% CI 0.05-1.34) that was sustained across 14-48 days (0.19, 95%CI 0.07-0.52) and 49-83 days (0.15, 0.07-0.32), with some apparent waning at 84+ days (0.47, 0.24-0.89) from receipt of booster dose. No hospitalisations were observed after second booster doses, but follow-up time was limited. Residents with known infection prior to the analysis period were at reduced risk of hospitalisation relative to those without prior infection, and within this group there were too few hospitalisation events to reliably estimate the effect of booster vaccination. Additional adjustment for primary vaccine course type did not improve model fit (P=0.72). Staff were at low risk of hospitalisation following SARS-CoV-2 infection, precluding meaningful analysis of the effect of booster vaccination.

### Death

In residents without known prior SARS-CoV-2 infection, the first booster reduced risk of death within 28 days of SARS-CoV-2 infection after 0-13 days (HR 0.14, 95%CI 0.02-1.04), 14-48 days (0.12, 0.04-0.34), and 49-83 days (0.11, 0.05-0.23), with apparent waning in the level of protection 84+ days (0.37, 0.21-0.62). Residents with known infection prior to the analysis period were at reduced risk of death relative to those without prior infection, and within this group there were too few deaths to evaluate the impact of booster vaccination. No deaths were observed after second booster doses, but limited follow-up time was available. Additional adjustment for primary vaccine course type did not improve model fit (P=0.70). No deaths within 28 days of a positive SARS-CoV-2 test were observed among staff.

### Reinfections

The appearance and spread of the Omicron variant in late 2021 and early 2022 was associated with a peak in incidence of new SARS-CoV-2 infections that exceeded the previous highest recorded levels in both residents and staff (Figure 2). There was also a substantial rise in the incidence of reinfections detected. In line with our Cox models, residents with prior SARS-CoV-2 infection only displayed moderately lower incidence of new infection than exposure-naïve residents during this period (Figure 2c), and in staff the incidence of infection was unrelated to history of previous infection (Figure 2d).

**Figure 2.**
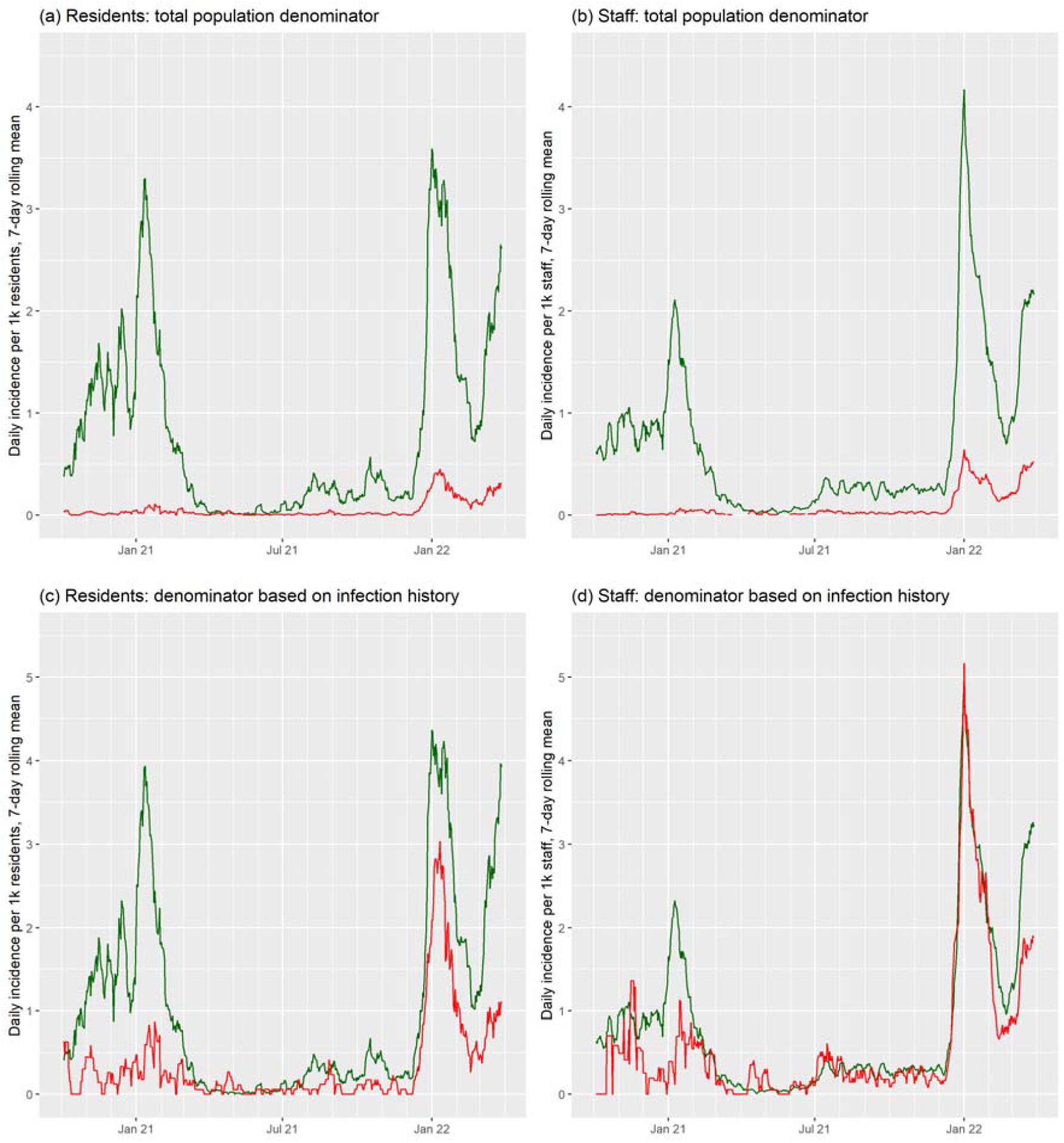
Rolling 7-day average incidence rate of new SARS-CoV-2 infections (green) and repeat SARS-CoV-2 infections (red) among residents and staff of long-term care facilities in the Vivaldi study. In parts (a) and (b), incidence rates are calculated according to the total population under follow-up for residents and staff (and are proportional to total case counts), whereas for (c) and (d) incidence rates are calculated using separate denominators for infection-naïve and infection-experienced participants (corresponding to individual risk).

Our analysis of the risk of infection in relation to booster vaccination only includes the first observed infection in any given participant within the analysis period. However, we observed two infections within the analysis period, using 30-day cut-off to define new infection episode, in 14 residents (0.10%) and 79 staff (0.40%).

## Discussion

We found that SARS-CoV-2 infection and associated hospitalisations and deaths in LTCF residents and staff who had received booster (third dose) vaccination were reduced compared to those who had only received primary vaccine course during the period of Omicron dominance in England. However, no protection against infection was apparent in residents from 84 days following third dose, and there was some waning of protection against hospitalisation and death. Whilst there was some evidence to suggest that infection rates were lower after second booster doses, the follow-up time was limited. Overall, these findings suggest that booster vaccination provides protection in residents against infection with the Omicron variant but that this protection wanes, with more moderate waning of protection against associated severe outcomes. Those with prior infection were susceptible to reinfection with Omicron.

Few studies have explored infection and severe outcomes after booster vaccination doses specifically in LTCFs and fewer still have explored this during the period in which the Omicron variant dominated. To the best of our knowledge, this is the only study to evaluate booster effectiveness in residents of LTCFs who have received AstraZeneca vaccine as primary course.

A Canadian study explored the effectiveness of fourth dose vaccination (primarily mRNA-1273), compared to third dose, among residents aged 60 years and older in LTCF and found evidence of improved protection against infection, symptomatic infection, and severe outcomes during the Omicron period[19]. A waning effect was observed after 84 days of third dose, but follow-up time after fourth dose was too limited for analysis[19]. A study conducted in the United States whilst Omicron was dominant estimated that VE against infection was 46.9% in LTCF residents who received a booster dose compared to those who had received 2 dose (primary course) vaccination, where booster vaccination was received 14 or more days prior to a positive test[20].

Our findings are also consistent with studies conducted in Israel which found that a third dose of the BNT162b2 mRNA vaccine compared with receipt of only two doses[21] and a fourth dose of the BN162b2 mRNA vaccine compared with three vaccine doses were effective in reducing risk of hospitalisation, severe disease and COVID-19 related death[22, 23]. There was evidence of waning effectiveness against infection[24], but sustained protection against severe disease[22, 23]. However, the study exploring third dose vaccine effectiveness was conducted in the general population and excluded healthcare staff and LTCF residents, while the fourth dose studies were conducted in older adults aged 60 years and over but not specifically LTCF residents. These findings may therefore not be generalisable to vulnerable residents of LTCFs.

This study has a number of strengths. Data were collected from a large cohort of LTCF residents and staff across England and linked to routinely collected and high-quality data on testing, hospitalisations, death and vaccination. The study population underwent regular, asymptomatic testing which enabled the systematic identification of study participants, accurate measurement of person-time at-risk and a comparatively unbiased assessment of vaccine effectiveness compared to studies that rely on symptomatic testing.

A limitation of the study is that it may have underestimated prior infection, as only a subset of participants underwent antibody testing. This may have resulted in an underestimate of the impact of past infection as people without antibody testing, along with those whose antibodies had waned, may have been misclassified as infection-naïve. Due to the nature of the data collection for hospitalisation and death records, it was not possible to distinguish between outcomes that occurred in individuals ‘with’ and ‘from’ COVID-19. Although our analysis focused on the period when the Omicron variant was dominant, we did not have access to sequencing data, so it was not possible to confirm viral sub-lineage or to investigate whether multiple positive PCR tests from the same individual over time were genuine reinfections. It was also not possible to include data on comorbidities which may impact on estimates of vaccine effectiveness.

This study suggests that third dose booster vaccination provides sustained protection against severe outcomes following infection with the Omicron variant in this vulnerable cohort, despite some waning, but protection against infection was not apparent from around 3 months onwards. In England, people aged over 75 years, including LTCF residents, and those who are clinically vulnerable are currently being offered a fourth COVID-19 vaccination dose[5]. Recent studies have shown fourth vaccine doses to be effective against severe illness caused by the Omicron variant when compared with a third dose administered 3-4 months previously in residents of LTCFs and older adults respectively[19, 23] but it is not yet known whether this pattern of waning immunity will continue to be seen. It seems likely that regular vaccination will be required for residents of LTCFs to ensure continued protection against SARS-CoV-2, particularly given the potential for the rapid emergence of new variants which may affect vaccine effectiveness. In this context, our findings underscore the critical need for continued surveillance of vaccine effectiveness against infection and severe outcomes in LTCFs, to inform future decisions on the frequency and timing of vaccination in this vulnerable cohort.

## Supporting information

STROBE checklist

## Data Availability

De-identified test results and limited meta-data will be made available for use by researchers in future studies, subject to appropriate research ethical approvals, once the VIVALDI study cohort has been finalised.

## Acknowledgements

We thank the staff and residents in the long-term care facilities who participated in this study and Mark Marshall at National Health Service (NHS) England who pseudonymised the electronic health records. This work is independent research funded by the Department of Health and Social Care (COVID-19 surveillance studies). MK is funded by a Wellcome Trust Clinical PhD Fellowship (222907/Z/21/Z). LS is funded by a National Institute for Health Research (NIHR) Clinician Scientist Award (CS-2016–007). AH is supported by Health Data Research UK (LOND1), which is funded by the UK Medical Research Council, Engineering and Physical Sciences Research Council, Economic and Social Research Council, Department of Health and Social Care (England), Chief Scientist Office of the Scottish Government Health and Social Care Directorates, Health and Social Care Research and Development Division (Welsh Government), Public Health Agency (Northern Ireland), British Heart Foundation, and Wellcome Trust. COVID-19 Genomics UK Consortium is supported by funding from the Medical Research Council (part of UK Research & Innovation), NIHR (grant code: MC_PC_19027), and Genome Research, operating as the Wellcome Sanger Institute. The views expressed in this publication are those of the authors and not necessarily those of the NHS, Public Health England, or the Department of Health and Social Care.

## Tables

**Table S1.** Characteristics of long-term care facilities of participants included in the analysis of booster vaccine effectiveness.

|  | **n (%) or median (IQR, range)** |
| --- | --- |
| Total | 328 |
| Capacity (median, IQR, range) | 5 (4-6, 1-17) |
| Facility type |  |
| For profit chain | 234 (71.3) |
| Residents | 9647 (68.1) |
| Staff | 12610 (63.7) |
| Capacity (median, IQR, range) | 50 (41-67, 5-169) |
| Not-for-profit chain | 67 (20.4) |
| Residents | 3445 (24.3) |
| Staff | 5494 (27.8) |
| Capacity (median, IQR, range) | 48 (40-60, 24-100) |
| Independent | 27 (8.2) |
| Residents | 1083 (7.6) |
| Staff | 1689 (8.5) |
| Capacity (median, IQR, range) | 41 (30-53, 22-90) |

**Table S2.** Crude event rates and adjusted hazard ratios against PCR or LFD-positive SARS-CoV2 infections for LTCF staff, by prior SARS-CoV2 exposure, primary vaccine course and booster vaccination status

| **SARS-CoV-2 infections: Staff with primary AstraZeneca course** |  |  |  |  |  |  |
| --- | --- | --- | --- | --- | --- | --- |
| **Prior SARS-CoV-2 exposure** | **Vaccination status** | **Person-days** | **Infections** | **IR /1000pd** | **HR (95% CI)\*** | **HR (95% CI)\*** |
| Unexposed | D2 84+d | 132680 | 508 | 3.83 | Ref. |  |
|  | D3 0-13d | 32131 | 77 | 2.4 | 0.26 (0.20-0.34) |  |
|  | D3 14-48d | 148200 | 271 | 1.83 | 0.28 (0.24-0.33) |  |
|  | D3 49-83d | 212023 | 390 | 1.84 | 0.46 (0.38-0.55) |  |
|  | D3 84+d | 295861 | 709 | 2.4 | 1.12 (0.97-1.29) |  |
|  | D4 0+d | 2410 | 2 | 0.83 | 0.52 (0.12-2.17) |  |
| Exposed | D2 84+d | 27967 | 106 | 3.79 | 1.05 (0.85-1.31) | Ref. |
|  | D3 0-13d | 4081 | 7 | 1.72 |  | 0.15 (0.07-0.32) |
|  | D3 14-48d | 16177 | 23 | 1.42 |  | 0.20 (0.12-0.33) |
|  | D3 49-83d | 27728 | 64 | 2.31 |  | 0.41 (0.29-0.58) |
|  | D3 84+d | 46600 | 113 | 2.42 |  | 1.08 (0.82-1.41) |
|  | D4 0+d | 229 | 0 | 0 |  | — |
| **SARS-CoV-2 infections: Staff with primary mRNA course** |  |  |  |  |  |  |
| **Prior SARS-CoV-2 exposure** | **Vaccination status** | **Person-days** | **Infections** | **IR /1000pd** | **HR (95% CI)\*** | **HR (95% CI) \*** |
| Unexposed | D2 84+d | 114645 | 435 | 3.79 | Ref. |  |
|  | D3 0-13d | 19664 | 70 | 3.56 | 0.31 (0.23-0.41) |  |
|  | D3 14-48d | 78281 | 174 | 2.22 | 0.37 (0.30-0.46) |  |
|  | D3 49-83d | 151090 | 363 | 2.4 | 0.43 (0.37-0.51) |  |
|  | D3 84+d | 350161 | 1041 | 2.97 | 1.26 (1.09-1.45) |  |
|  | D4 0+d | 2266 | 4 | 1.77 | 0.97 (0.35-2.64) |  |
| Exposed | D2 84+d | 15883 | 59 | 3.71 | 0.96 (0.75-1.25) | Ref. |
|  | D3 0-13d | 2370 | 3 | 1.27 |  | 0.09 (0.03-0.30) |
|  | D3 14-48d | 9013 | 13 | 1.44 |  | 0.25 (0.13-0.48) |
|  | D3 49-83d | 21479 | 55 | 2.56 |  | 0.44 (0.30-0.65) |
|  | D3 84+d | 61778 | 194 | 3.14 |  | 1.36 (1.01-1.83) |
|  | D4 0+d | 189 | 0 | 0 |  | — |

\*HR values in the two columns are from mathematically identical statistical models, but HRs in right-hand column are expressed relative to 'D2 84+d' vaccine status in individuals with prior SARS-CoV-2 infection.

